# Rheumatoid arthritis and idiopathic pulmonary fibrosis: a bidirectional Mendelian randomisation study

**DOI:** 10.1101/2022.09.27.22280286

**Authors:** Olivia C Leavy, Leticia Kawano-Dourado, Iain D Stewart, Jennifer K Quint, Joshua J Solomon, Raphael Borie, Bruno Crestani, Louise V Wain, R Gisli Jenkins, Philippe Dieudé, Cosetta Minelli

## Abstract

**Introduction:** A usual interstitial pneumonia (UIP) pattern of lung injury is a key feature of idiopathic pulmonary fibrosis (IPF) and is also observed in up to 40% of individuals with rheumatoid arthritis (RA) related Interstitial Lung Disease (RA-ILD). The RA-UIP phenotype could result from either a causal relationship of RA on UIP or vice versa, or from a simple co-occurrence of RA and IPF due to shared demographic, genetic or environmental risk factors.

**Methods:** We used two-sample bidirectional Mendelian Randomisation (MR) to investigate the causal effects of RA on UIP and of UIP on RA, using variants from genome-wide association studies of RA (separately for seropositive and seronegative RA) and of IPF as genetic instruments. We conducted inverse-variance-weighted fixed-effect MR as a primary analysis and undertook sensitivity analyses to assess potential violations of the key MR assumption of no (horizontal) pleiotropy.

**Results:** Seropositive RA showed a significant protective effect on IPF (Odds Ratio, OR = 0.93; 95% Confidence Interval, CI: 0.87-0.99; *P*=0.032), while the MR in the other direction showed a strongly significant causal effect of IPF on seropositive RA (OR = 1.06, 95% CI: 1.04-1.08, *P*=1.22×10^−11^).

**Conclusion:** Our findings support the hypothesis that RA-UIP may be due to a cause-effect relationship between IPF and RA, rather than due to a coincidental occurrence of IPF in patients with RA. The causal effect of IPF on seropositive RA suggests that patho-mechanisms involved in the development of UIP may promote RA and would suggest that screening for UIP in asymptomatic RA patients may be warranted and may influence therapy management of patients with RA-UIP.

## Introduction

Interstitial Lung Disease (ILD) is common in Rheumatoid Arthritis (RA) and is considered an extra-articular manifestation of RA that may occur at different stages of the disease and significantly influences prognosis (1–5). Though several histopathological/high-resolution computerized tomography (HRCT) patterns of lung damage in RA-ILD have been described, usual interstitial pneumonia (UIP) is the most frequent, seen in 40% of individuals with RA-ILD (6) (where it is termed RA-UIP), followed by nonspecific interstitial pneumonia (NSIP) which is seen in 30% (1–5, 7).

UIP is also the histopathological pattern of idiopathic pulmonary fibrosis (IPF), a progressive and fatal scarring disease of the lungs. RA-UIP and IPF share several clinical features such as a male sex predominance, older age at onset (around the 6^th^ and 7^th^ decade, respectively), indistinguishable patterns of ILD on HRCT, a poor prognosis (8–13) and a similar magnitude of response to anti-fibrotic therapy (14, 15).

RA-UIP and IPF also share risk factors, including smoking and the *MUC5B* rs35705950 genetic variant (T risk allele), suggesting common pathogenic pathways (16–18). Indeed, the association between RA-ILD and *MUC5B* rs35705950 is restricted to the RA-UIP subtype of RA-ILD with a similar magnitude and direction to that reported in IPF (16, 19). Of note, *MUC5B* rs35705950 was not found to contribute to the risk of RA without ILD (16). However, provocatively, the restricted association of *MUC5B* rs35705950 with RA and a UIP pattern of injury (but not NSIP) raises the hypothesis that RA-UIP might in fact be a coincidental occurrence of IPF in individuals who also have RA, rather than UIP being a direct consequence of RA (20).

Observational studies cannot provide strong evidence on causal relationships, nor the direction of causation, as they are vulnerable to confounding and reverse causation. Mendelian Randomisation (MR) is a statistical approach that can infer causal relationships between two traits and the direction of causality using genetic variants as instrumental variables (IVs). MR can be considered as a “natural” randomised control trial as the genetic variants an individual holds are randomly assigned at conception and do not vary during their lifetime, thus being not subject to confounding or reverse causation. For an MR study to be valid three key assumptions about the IVs should hold; 1) they should be associated with the exposure (risk factor) of interest, 2) they should not be associated with confounders of the exposure and outcome relationship, and 3) they should not be associated with the outcome other than through the exposure (i.e., no horizontal pleiotropy). MR results can be biased by horizontal pleiotropy, but there are methods available to detect and allow for pleiotropy.

We hypothesised that the RA-UIP phenotype was a simple co-occurrence of RA and IPF, rather than the result of either a causal relationship of RA on UIP or vice versa. To test this, we undertook a MR analysis to estimate the causal relationship between RA and IPF. We used a two-sample approach where we derived causal estimates from separate studies of RA and IPF. Our analysis was bidirectional, testing both for a causal effect of RA on IPF, and for a causal effect of IPF on RA. We undertook separate analyses for seropositive and seronegative (for Rheumatoid Factor (RF) and/or anti-citrullinated protein/peptide antibody (ACPA)) RA.

## Methods

For our bidirectional MR analysis, we used a two-sample approach where summary statistics (i.e. effect estimates and standard errors) for the gene-exposure (“G-X”) and gene-outcome (“G-Y”) associations were obtained from separate studies. For the MR of the effect of RA on IPF, G-X refers to genetic associations with RA and G-Y to genetic associations with IPF, and vice versa for the MR of IPF on RA (Figure 1).

**Figure 1:**
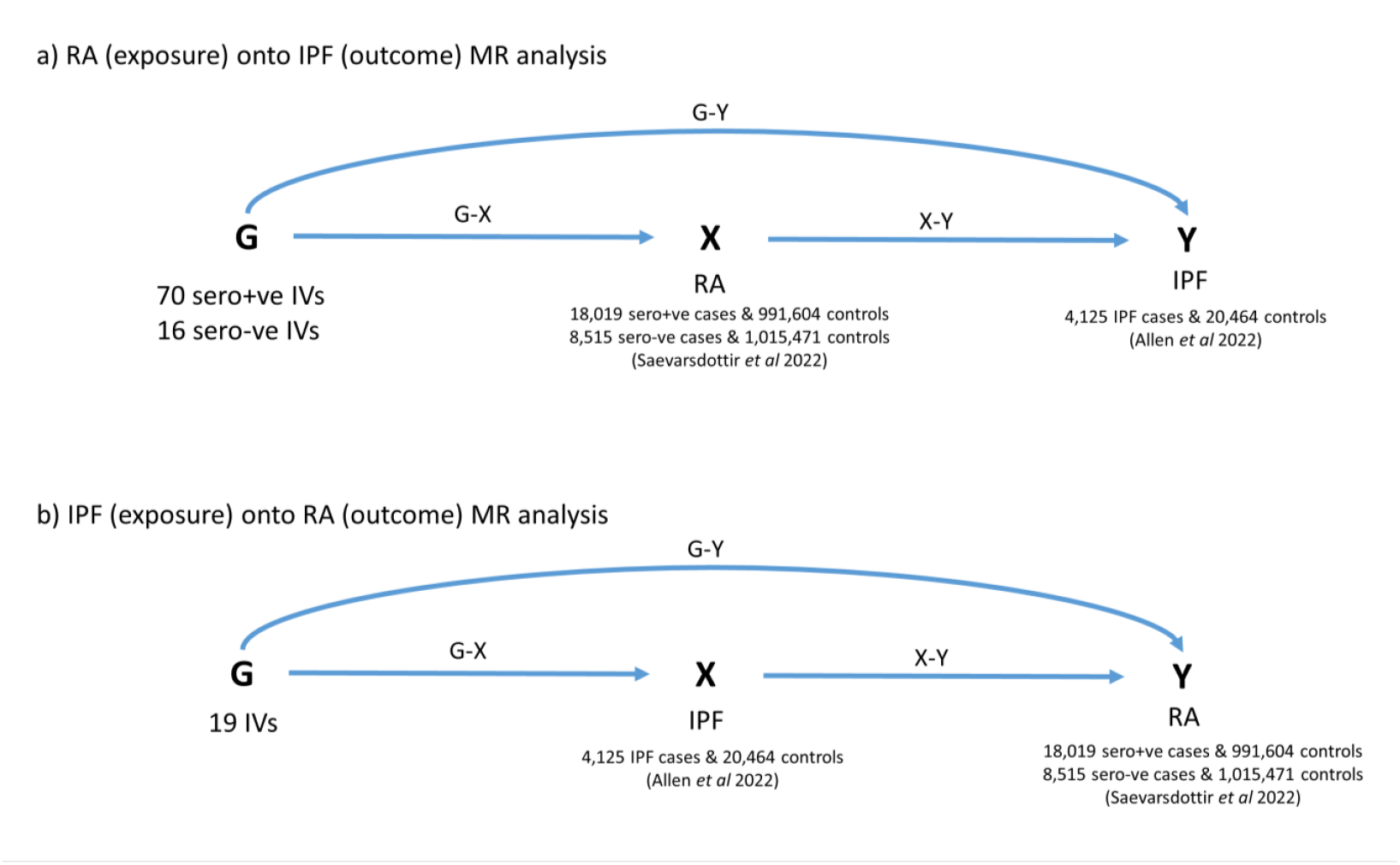
Directed acyclic graphs illustrating MR analyses and the number of instrumental variables used for a) RA (exposure) onto IPF (outcome) MR analysis and b) IPF (exposure) onto RA (outcome) MR analysis

### Study populations

Genetic association estimates for seropositive and seronegative RA were taken from a published study of RA (21) that reported separate genome-wide association studies (GWAS) of seropositive (18,019 cases and 991,604 controls) and seronegative (8,515 cases and 1,015,471 controls) RA. One hundred and thirty-five autosomal single nucleotide polymorphisms (SNPs) that were associated with RA (either seropositive, seronegative or both) in previous GWAS of European ancestry or multi-ancestry were selected as IVs for RA (22). These IVs were used for testing the causal effect of RA as the exposure and on IPF as the outcome.

Genetic association estimates for IPF were obtained from a previously published GWAS comprising 4,125 IPF cases and 20,464 controls (23). Nineteen common SNPs reported as being genome-wide significantly associated with IPF were selected as IVs for IPF (23). These IVs were used for testing the causal effect of IPF as the exposure and on RA as the outcome.

SNPs were excluded from the analyses if they were not present in the relevant outcome dataset and no suitable proxy (linkage disequilibrium r^2^>0.8) could be found. For palindromic SNPs (i.e., A/T, C/G SNPs), non-palindromic proxies (r^2^>0.8) were selected. For correlated SNPs (r^2^>0.01, determined using 1000 Genomes Project EUR population using LDlink (24)), the SNP with the least significant association with the exposure was excluded.

As the use of “weak” instruments can bias the results of MR, SNPs with an F-statistic < 10 were excluded, where the F-statistic represents a measure of instrument strength (25, 26).

### Statistical analyses

The inverse-variance weighted (27) fixed effect (IVW-FE) method is a fixed-effect meta-analysis of MR estimates across SNPs, where SNP-specific MR estimates are obtained using the Wald estimator (G-Y/G-X). This was used for the primary analysis for the MR in both directions, as it is the most powerful MR method in the absence of pleiotropy.

To investigate presence and magnitude of pleiotropy, the Cochran’s Q statistic and I^2^ statistic were used, respectively. Individual variant contributions to Cochran’s Q heterogeneity statistic were plotted to identify pleiotropic SNPs (28). In the presence of pleiotropy, a series of sensitivity analyses were conducted to account for it; inverse-variance weighted random-effect (IVW-RE) method, MR-PRESSO (MR Pleiotropy RESidual Sum and Outlier) (29), weighted median (30), weighted mode-based estimation (31) and MR-Egger (32). These different methods perform better in different scenarios as they make different assumptions about the nature of the underlying pleiotropy.

IVW-RE is an inverse-variance weighted method where the fixed-effect meta-analysis model of IVW-FE is substituted by a random-effects model to allow for heterogeneity (as a proxy for pleiotropy). In particular, this method allows for balanced pleiotropy (random effects have a mean of zero), in the presence of which the point estimate is equivalent to the IVW-FE point estimate, but IVW-RE will have wider 95% CIs.

MR-Egger allows all SNPs to have pleiotropic effects, however pleiotropic effects should be independent of the G-X associations. The method is affected by outliers, particularly when the G-X estimates are similar across different SNP, which in turn can cause there to be low power to detect a causal effect. When the variation in the strength of the instruments is limited, MR-Egger is susceptible to dilution bias, which biases the MR results towards the null. The I^2^ of a meta-analysis of G-X estimates (I^2^_GX_) can be used to assess this, with lower values suggesting stronger dilution. Ideally, the I^2^_GX_ measure should be >90%; when this is not the case, MR-Egger should be performed using simulation extrapolation (SIMEX) to correct for the dilution bias (33).The weighted median method makes weaker assumptions about valid IVs, as it only assumes that at least half of the variants are valid instruments. This method is robust to outliers and is not as affected by the presence of a small number of pleiotropic variants as the IVW and MR-Egger methods.

The weighted mode method is also robust to outliers and it makes even weaker assumptions, only assuming that the largest (weighted) contribution of similar SNP-specific MR estimates comes from valid instruments.

MR-PRESSO can be used to identify and remove possible pleiotropic SNPs which have been detected in the MR analysis as outliers. However, the outlier test requires at least 50% of the genetic variants used as valid IVs.

We also performed a leave-one-out analysis, where each IV is excluded in turn and the analysis repeated to identify whether the results are highly influenced by a single IV, to determine whether any of the causal estimates were heavily influenced by individual instruments.

All analyses were performed using packages in R (version 4.1.0), specifically “*MendelianRandomization*” (for IVW-FE, IVW-RE, MR-Egger, weighted mode and weighted median), “*MRPRESSO*” (for MR-PRESSO), “*simex*” (for MR-Egger with SIMEX extension) and “*TwoSampleMR*” (to harmonise the RA and IPF summary data and to perform the leave-one-out analyses). Estimates for Cochran’s Q test and I^2^ were obtained using IVW-FE analysis.

## Results

### Selection of genetic instruments

Of the 135 IVs initially selected as IVs for RA, two were associated with seronegative RA only, one was associated with seronegative and combined RA, 93 were associated with combined RA and 39 were associated with seropositive RA. For the seropositive analysis we selected IVs associated with seropositive RA or combined RA and for the seronegative analysis IVs associated with seronegative RA or combined RA. In total, 70 were strong instruments (F-statistic >=10) for seropositive RA (Supplementary Table S1) and 16 were strong instruments for seronegative RA (Supplementary Table S2). For IPF, all 19 association signals reported by Allen *et al* 2022 were strong instruments for IPF (Supplementary Tables S3 and S4).

### Causal estimate for seropositive RA on IPF

The primary IVW-FE analysis gave a nominally significant result for a protective causal effect of seropositive RA on IPF (odds ratio (OR) 0.93; 95% confidence interval (CI) 0.87-0.99; *p*=0.032) (Figure 2a and Supplementary Table S5a). Although there was statistically significant evidence of pleiotropy (Q test *p*-value = 2×10^−4^, and MR-PRESSO global test *p*-value = 2×10^−4^), this was of moderate magnitude (I^2^ = 41.3%, 95% CI = 22%-56%) and no SNPs were highlighted as outliers when using MR-PRESSO or when plotting individual contributions to Cochran’s Q heterogeneity (Supplementary Figure S1). Moreover, significant estimates of a protective causal effect of seropositive RA on IPF were also obtained using the weighted median, weighted mode and MR-Egger analyses, and in the leave-one-out analysis, no exclusions resulted in a change in the direction of effect (Supplementary Figure S2).

**Figure 2:**
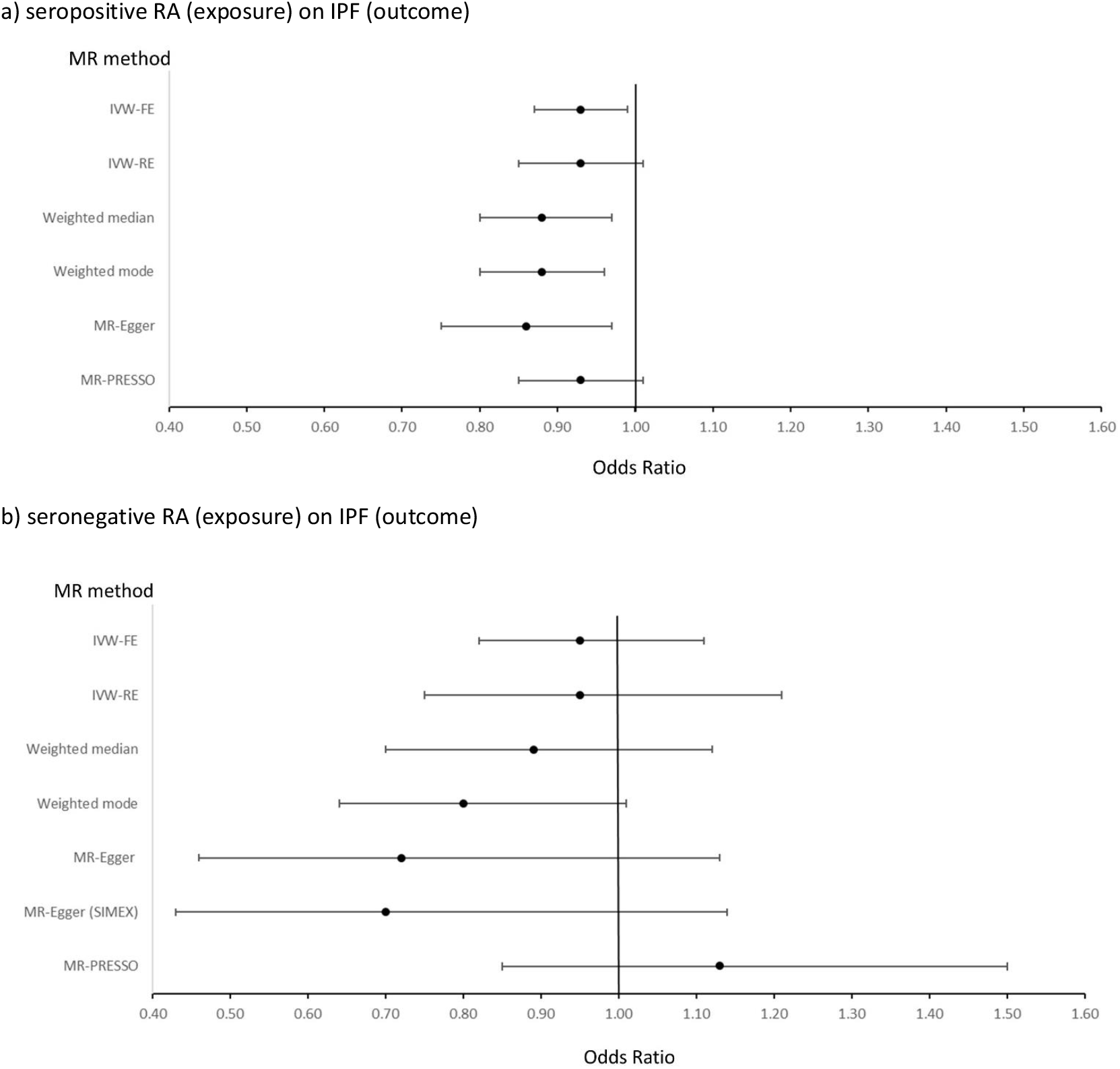
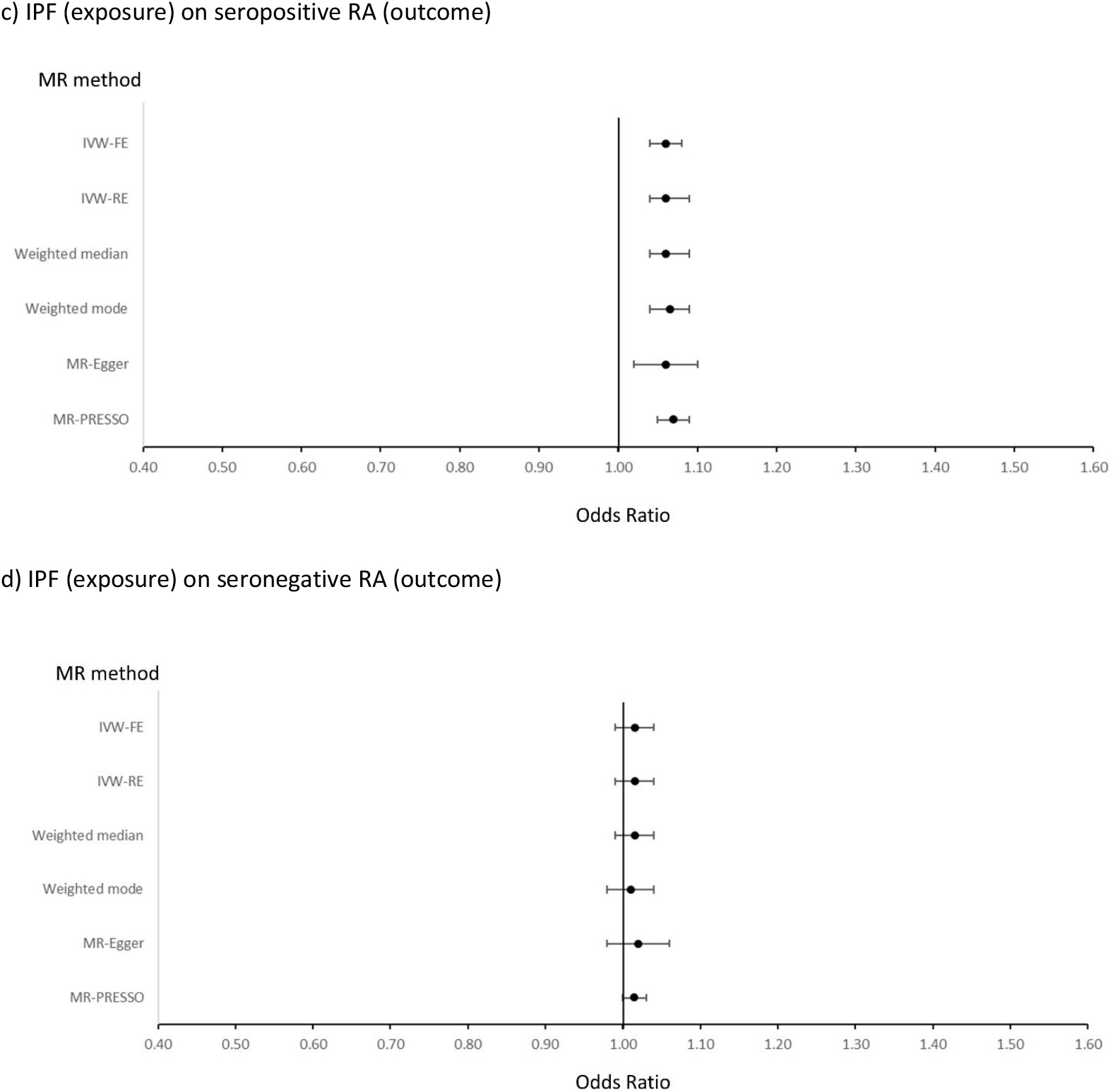
Forest plots of results of Mendelian Randomisation to estimate the causal effect of a) seropositive RA (exposure) on IPF (outcome), b) seronegative RA (exposure) on IPF (outcome), c) IPF (exposure) on seropositive RA (outcome) and d) IPF (exposure) on seronegative RA (outcome)

### Causal estimate for seronegative RA on IPF

Whilst the IVW-FE point estimate was similar to that of seropositive RA, the confidence intervals were very wide and the result was non-significant (95% CI 0.82-1.11, *p*=0.556) (Figure 2b and Supplementary Table S5a). The SNP rs6910071 was identified as an outlier by MR-PRESSO and there was statistically significant evidence of pleiotropy (Q test *p*-value = 0.002, and MR-PRESSO global test *p*-value = 0.0087, I^2^ = 57.8%, I^2^ 95% CI = 27%-76%). Even after removing the SNP rs6910071 (positioned in the HLA region of chromosome 6) from the MR analysis (leave-one out analysis), the results remained null (Supplementary Figure S3).

### Causal estimate for IPF on seropositive RA

The primary IVW-FE analysis suggested that developing IPF increases the risk of seropositive RA (OR 1.06, 95% CI 1.04-1.08, *p*=1.22×10^−11^) (Figure 2c and Supplementary Table S5b). There was significant evidence of pleiotropy (Q test *p*-value = 0.008, and MR-PRESSO global test *p*-value = 0.0313; I^2^ = 49.4%, I^2^ 95% CI = 14%-70%). Two outliers were detected and removed by MR-PRESSO (rs112087793 and rs12912339), however, the causal effect of IPF on seropositive RA remained significant (OR 1.07, 95% CI 1.05-1.09, *p*=1.27×10^−7^). The point estimates for the IVW-FE, weighted median, weighted mode and MR-Egger were also consistent and all results were statistically significant.

In the leave one out analyses, no individual variant exclusions substantially attenuated the original result (Supplementary Figure S4).

### Causal estimate for IPF on seronegative RA

None of the analyses suggested a significant causal effect of IPF on seronegative RA (Figure 2d and Supplementary Table S5b) although all point estimates were greater than one, consistent with the seropositive analysis. There was no evidence of pleiotropy, MR-PRESSO did not detect any outliers and the leave-one-out analysis did not suggest that the result was influenced by any single IV (Supplementary Figure S5).

## Discussion

This bidirectional two-sample MR study did not support the hypothesis that RA-UIP is a coincidental occurrence of IPF in patients with RA, and instead provides evidence of a causal effect of IPF on the development of seropositive RA. There was also evidence for a protective causal effect of seropositive RA on development of IPF, albeit this was considerably weaker.

The rationale for a causal effect of RA on UIP has been driven by the temporal relationship between the two conditions with RA often being diagnosed before pulmonary fibrosis. However, several arguments could suggest a causal effect of IPF on RA. The loss of immune tolerance that occurs when the lungs are chronically damaged may suggest that IPF could be a risk factor for RA, as suggested by RA developing after IPF diagnosis (34). An independent study, the Multi-Ethnic Study of Atherosclerosis, measured RA-related auto-antibodies and obtained cardiac CT scans which were assessed for subclinical ILD. This study demonstrated an association between elevated RF, ACPA and subclinical ILD, suggesting autoantibody production and pulmonary inflammation develop prior to clinical RA (35). However, as it was unknown if participants had RA, this may represent an association of antibodies with subclinical ILD. An assessment of patients with ILD who did not fulfil the classification criteria for RA revealed a third of patients who were ACPA-positive developed RA less than three years of their ILD diagnosis (36). Furthermore, there is a significant proportion of patients with RA (27% to 48%) for whom the diagnosis of ILD precedes or occurs at the same time as the onset of RA (5, 37, 38). Of note, a majority of individuals who developed ILD prior to RA were found to have a radiological UIP pattern (37). Finally, it has been shown that IgA-ACPA are elevated in up to 25% of patients with IPF and associated with changes in pathology (*i*.*e*., lymphoid aggregates) also seen in established RA-UIP (39).

The mechanism by which pulmonary fibrosis may promote RA is likely *via* breaching immune tolerance against citrullinated peptides. Indeed, several indirect arguments have led to the hypothesis of a mucosal origin of seropositive RA (40), positioning the lung as the site of initiation of the loss-of-tolerance against citrullinated peptides; 1) most of the environmental risk actors in seropositive RA are inhaled (smoking, silica exposure), 2) in patients with early ACPA positivity (up to 15 years before the onset of the first joint manifestations)(41), the IgA isotype, predominates (42), 3) the presence of peptidyl arginine deiminase 2 (PAD2) in lung tissue, an enzyme responsible for citrullination, and local production of ACPA (43–45), and 4) the existence of shared citrullinated peptide targets in lungs and joints of RA patients (46). These data suggest the possibility that in a subset of patients, the citrullinated protein targets of ACPA are lung-specific, leading to lung injury and fibrosis and, through a broadening of the ACPA repertoire, eventual synovitis and clinical RA (47, 48).

Mucosal inflammation has long been considered the source of ACPA associated with seropositive RA, particularly IgA isotypes, and lymphoid follicles are common in both IPF and RA (40, 45). It is thought that chronic infection or changes in the microbiome can promote protein citrullination via chronic inflammation and NETosis (49, 50). Indeed, the predominant microbiota of both IPF and RA has been found to be the phyla Firmicutes (51–54).

A fundamental principle of clinical management is to treat the underlying cause of any disease. Therefore, understanding the direction of causality between two associated conditions is crucial. RA has been considered causal for UIP for many years and has guided therapeutic decisions such as prioritising the use of immunomodulatory therapy in patients with RA-UIP. Our data would suggest re-evaluating the therapeutic paradigm for treating RA-UIP. Indeed, the first randomised, double-blind, placebo-controlled trial dedicated to patients with RA-ILD identified that pirfenidone had a greater effect on slowing the decline of FVC in patients with a UIP pattern on HRCT (14). Secondly, our findings raise the intriguing hypothesis that early identification and treatment of UIP in patients with RA may offer a novel strategy for managing RA. Historically, rheumatologists have been reluctant to screen asymptomatic patients with RA for ILD at point of diagnosis and these results would suggest that this approach should be reconsidered. Lastly, our findings also suggests that pulmonologists should carefully follow the outcome of patients with IPF, paying attention to the apparition of a RA specific autoimmunity as well as articular manifestations.

Whilst MR provides a framework to assess causality by using genetic instruments to remove the effects of confounders and reverse causation, it does have limitations. Possible pleiotropy was detected in most of our MR analyses, although the consistency of the results across different methods allowing for pleiotropy suggests robustness of our findings. The limited number of IVs and sample size of the source GWAS that were used to derive the causal estimates for IPF and seronegative RA, compared with the seropositive RA GWAS, may have impacted power to detect a causal relationship. Our hypothesis motivating this study was that RA-UIP is actually a co-occurrence of RA and IPF in the same patient, and so we used SNP IVs derived from IPF GWAS to model this.

Having detected a causal relationship between IPF and seropositive RA, it would be relevant to understand whether the causal effect was due to mechanisms promoting the UIP pattern of lung damage. However, no GWAS specifically for UIP exists and so the IPF SNP IVs are the best proxies for UIP IVs at this time. As the presence of UIP in the RA cases included in the RA source GWAS cannot be excluded, for the MR on the effect of RA on IPF, it is possible that some of the RA SNP IVs (“G-X”) could be specifically associated with RA-UIP. These SNPs might also be associated with IPF (UIP) in which case they could introduce a bias towards a causal effect of RA on IPF; however, this was not seen. As for the MR on the effect of IPF on RA, it is also possible that RA-UIP cases in the RA source GWAS might lead to an association with the IPF SNP IVs (“G-Y”) thereby introducing a bias towards a causal effect of IPF on RA. However, our leave-one-out analyses show that excluding the *MUC5B* SNP rs35705950, which is known to be associated with RA-UIP (16) with an effect that is similar in magnitude to the effect on IPF, did not change the causal effect estimate (Supplementary Figure S4). Whilst the direction and significance of the point estimates support a causal effect of IPF on seropositive RA, the magnitude of the effect should be interpreted with caution (55).

In spite of these limitations, our data suggest that patho-mechanisms involved in the development of UIP may promote RA. This has implications for the management of patients with RA-UIP. In addition, the causal effect of IPF on the development of seropositive RA would provide additional support for screening for ILD in patients with RA, especially in subgroups of patients identified as being at high-risk for pulmonary fibrosis (56, 57). Whilst we found no support for a causal effect of RA on IPF, the opposite finding of a significant protective effect of RA against development of IPF was unexpected and requires further investigation.

## Data Availability

All data produced are available online at:

https://github.com/genomicsITER/PFgenetics

https://www.decode.com/summarydata/

## Data Availability

IPF GWAS data are available from: https://github.com/genomicsITER/PFgenetics

RA GWAS data are available from: https://www.decode.com/summarydata/

## Funding and acknowledgements

LVW holds a GSK / Asthma + Lung UK Chair in Respiratory Research (C17-1). The research was partially supported by the National Institute for Health Research (NIHR) Leicester Biomedical Research Centre; the views expressed are those of the author(s) and not necessarily those of the National Health Service (NHS), the NIHR or the Department of Health. This research used the SPECTRE High Performance Computing Facility at the University of Leicester. LVW, JKQ and RGJ were supported by the Medical Research Council (MR/W014491/1, DEMISTIFI). For the purpose of open access, a CC BY or equivalent licence will be applied to any author accepted manuscript version arising from this submission.

## Supplementary Tables/Figures

Supplementary Table S1: Seropositive RA and IPF association results, F-statistics and MR causal effects for seropositive RA instrumental variables

[Excel spreadsheet: Supplementary Tables]

Supplementary Table S2: Seronegative RA and IPF association results, F-statistics and MR causal effects for seronegative RA instrumental variables

[Excel spreadsheet: Supplementary Tables]

Supplementary Table S3: IPF and seropositive RA association results, F-statistics and MR causal effects for IPF instrumental variables

[Excel spreadsheet: Supplementary Tables]

Supplementary Table S4: IPF and seronegative RA association results, F-statistics and MR causal effects for IPF instrumental variables

[Excel spreadsheet: Supplementary Tables]

**Supplementary Table S5:**
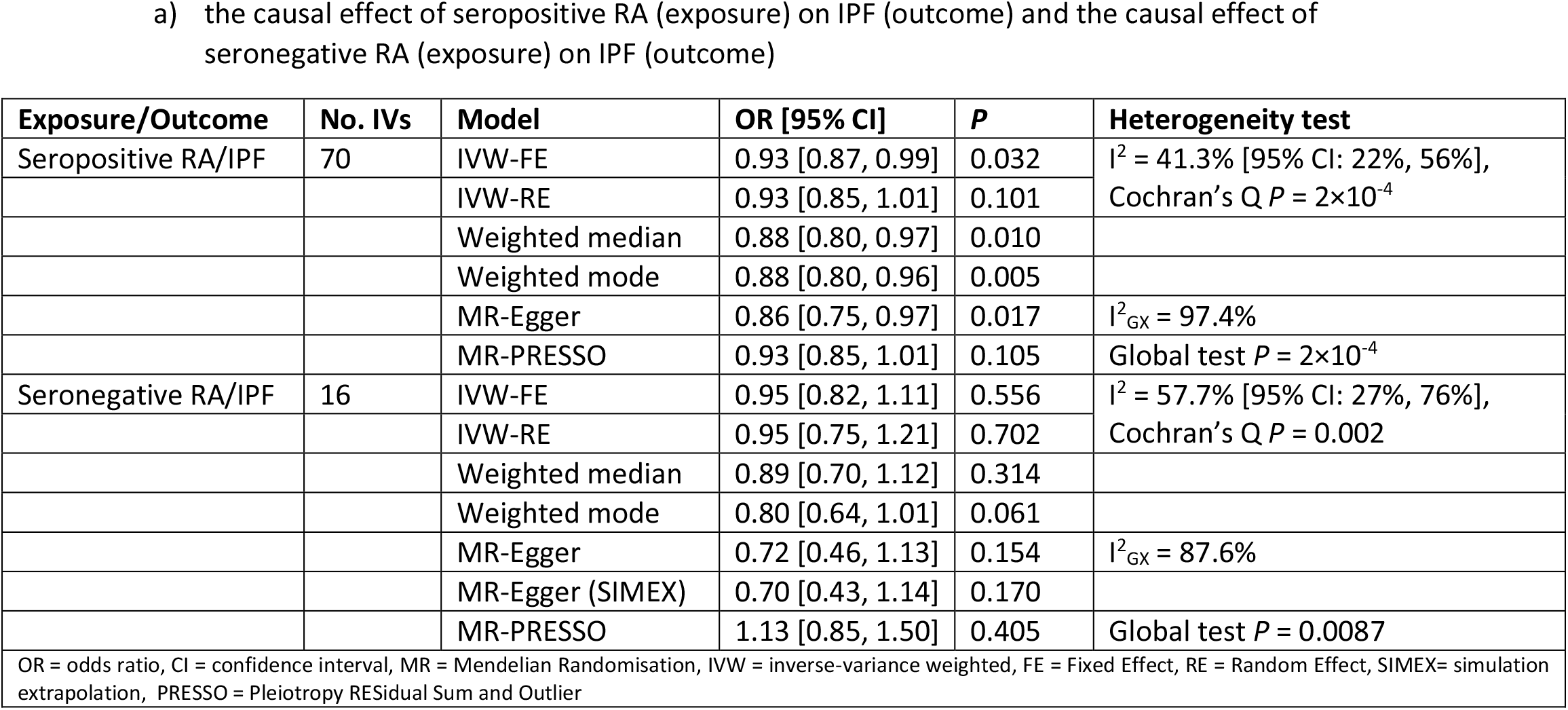
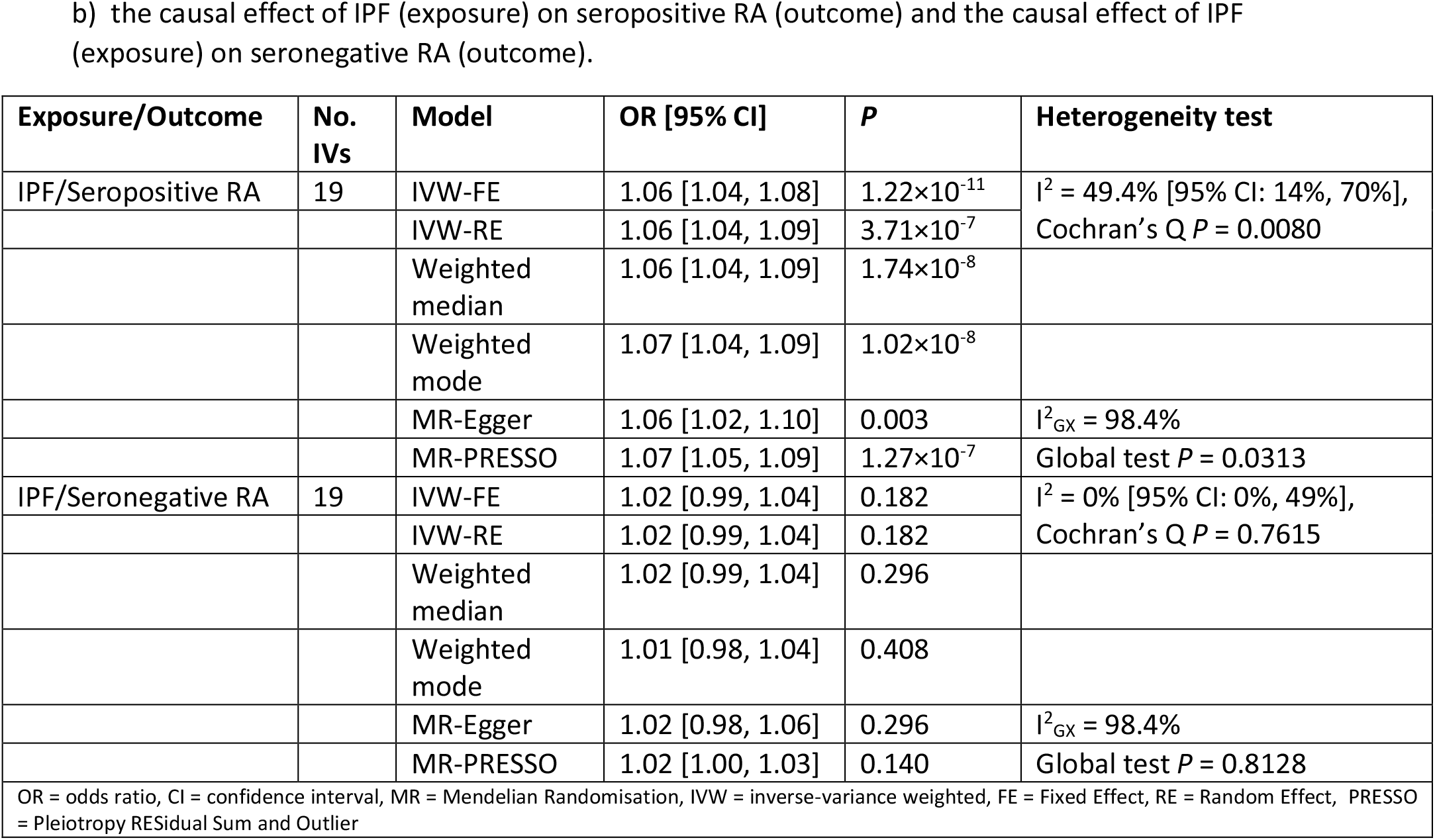
Results of Mendelian Randomisation analyses to estimate the causal effect of a) seropositive RA (exposure) on IPF (outcome) and the causal effect of seronegative RA (exposure) on IPF (outcome), b) the causal effect of IPF (exposure) on seropositive RA (outcome) and the causal effect of IPF (exposure) on seronegative RA (outcome).

**Supplementary Figure S1:**
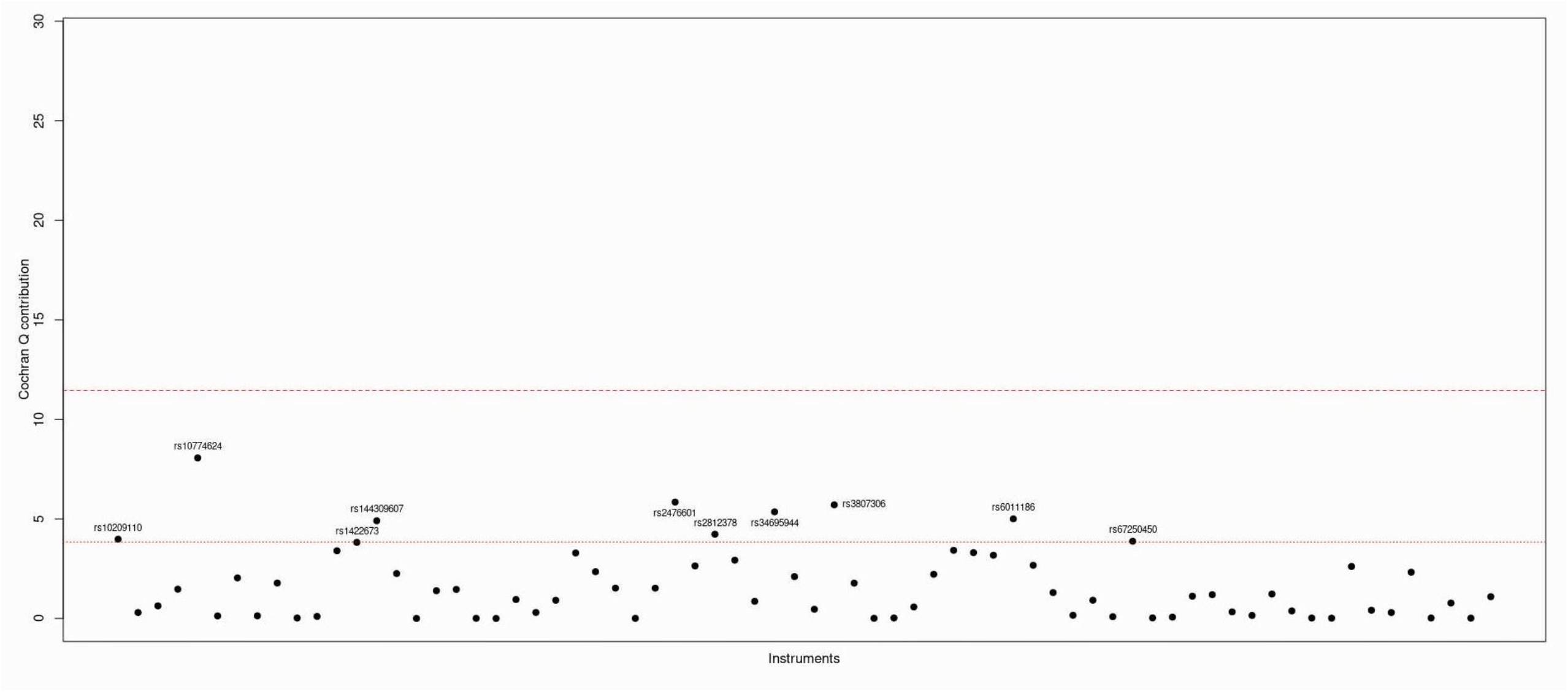
Plot of Cochran Q contribution for each seropositive RA instrumental variable

**Supplementary Figure S2:**
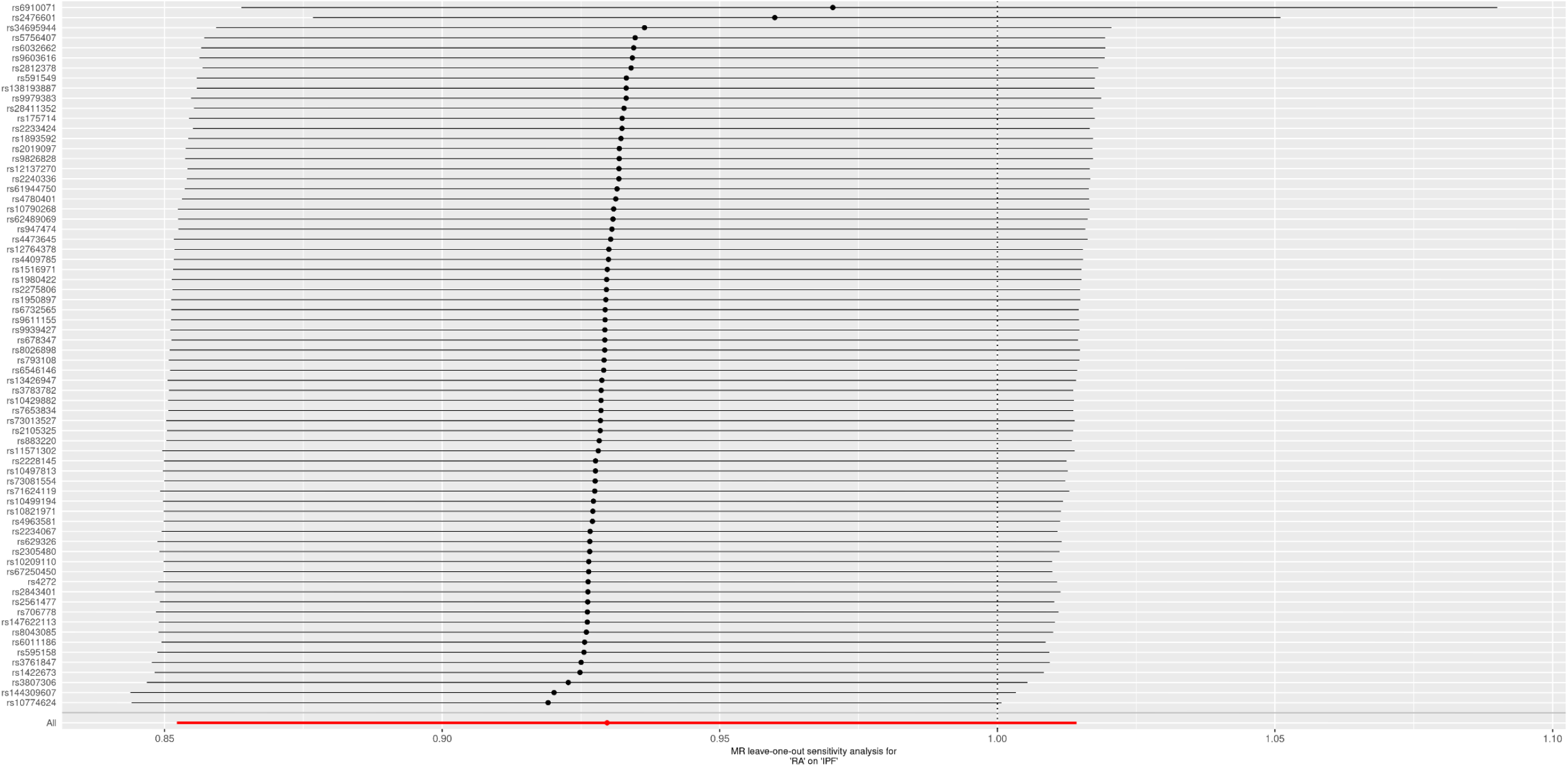
Leave one out analysis for estimation of causal effect of seropositive RA on IPF

**Supplementary Figure S3:**
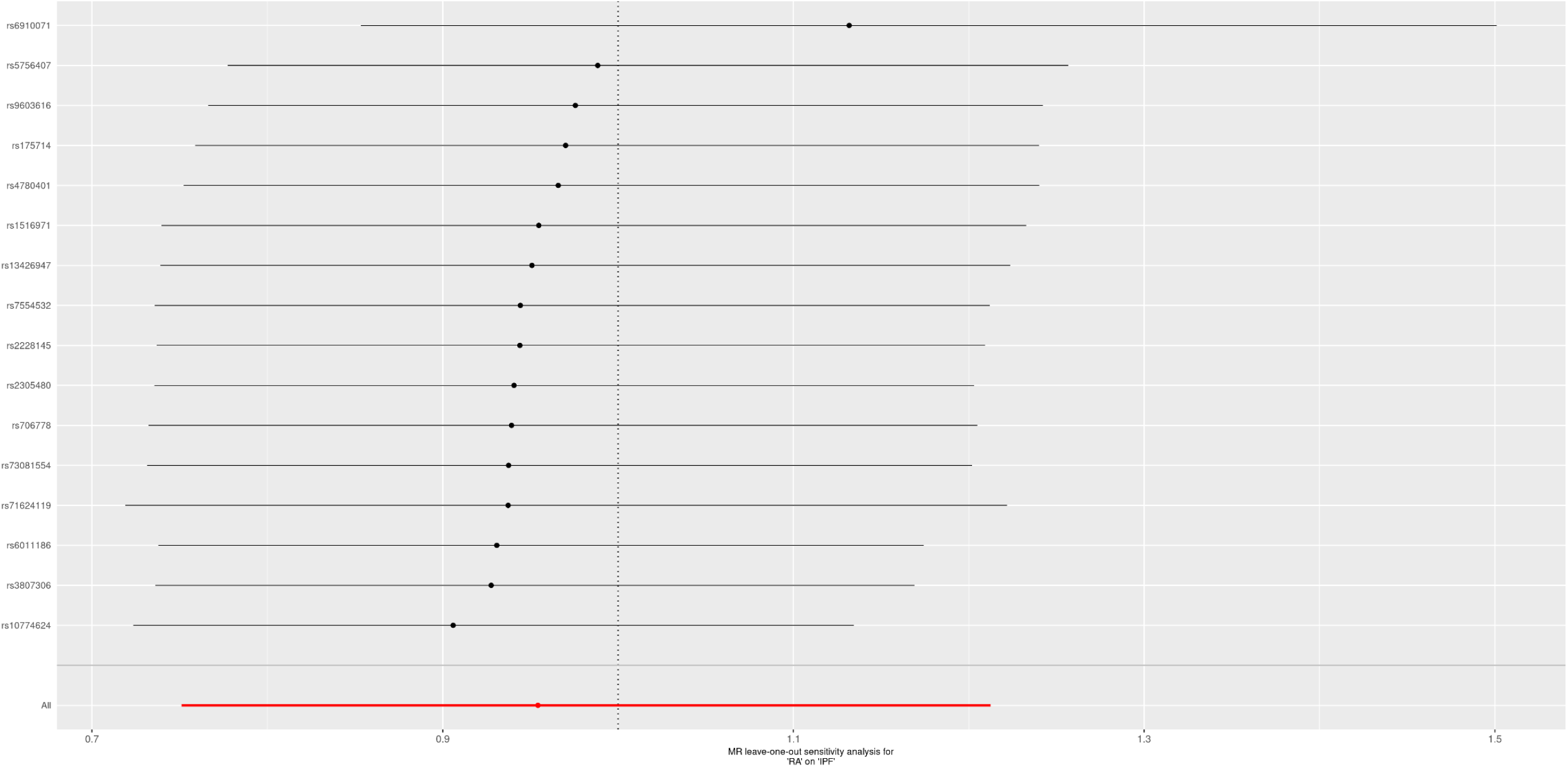
Leave one out analysis for estimation of causal effect of seronegative RA on IPF

**Supplementary Figure S4:**
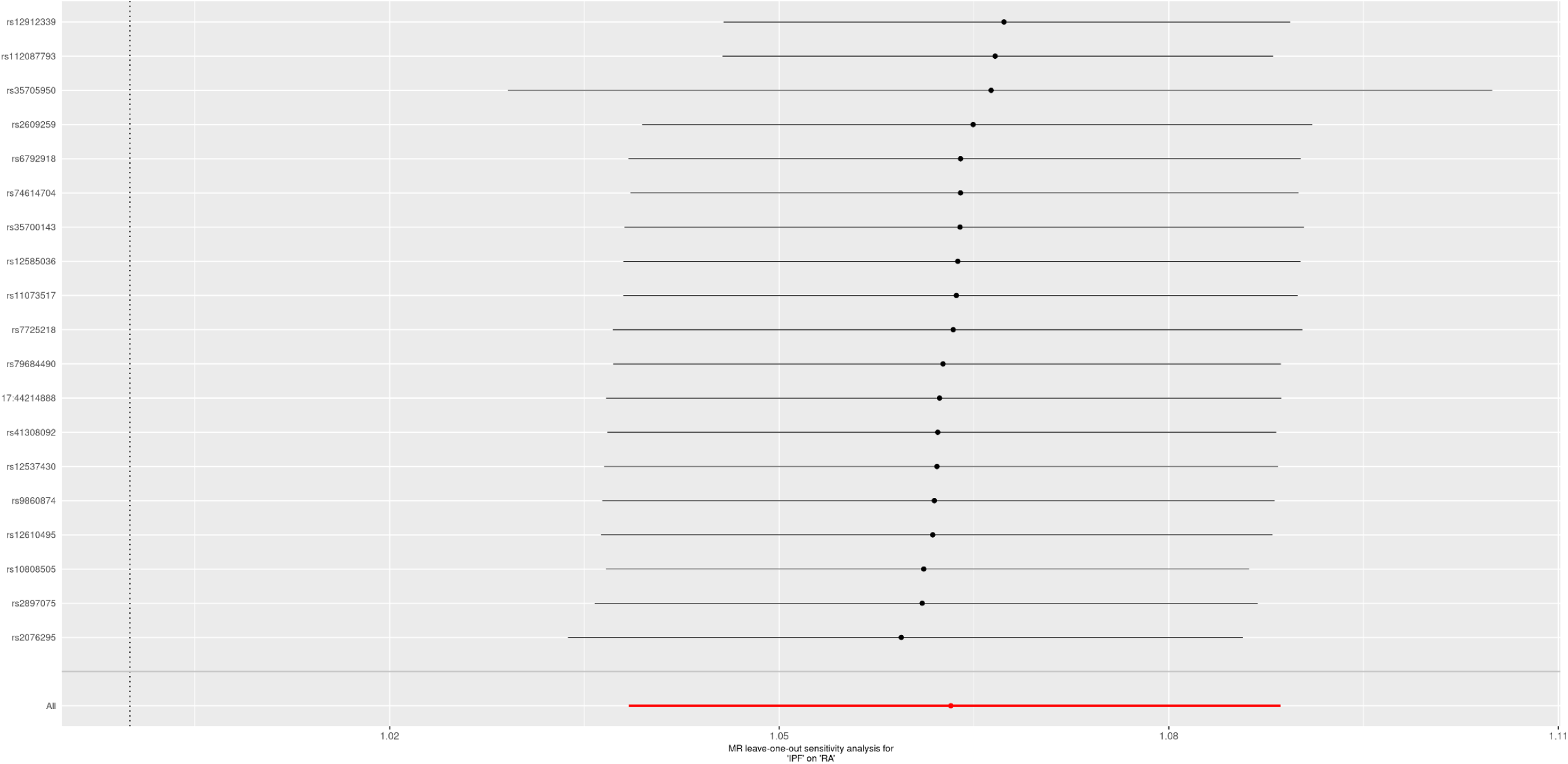
Leave one out analysis for estimation of causal effect of IPF on seropositive RA

**Supplementary Figure S5:**
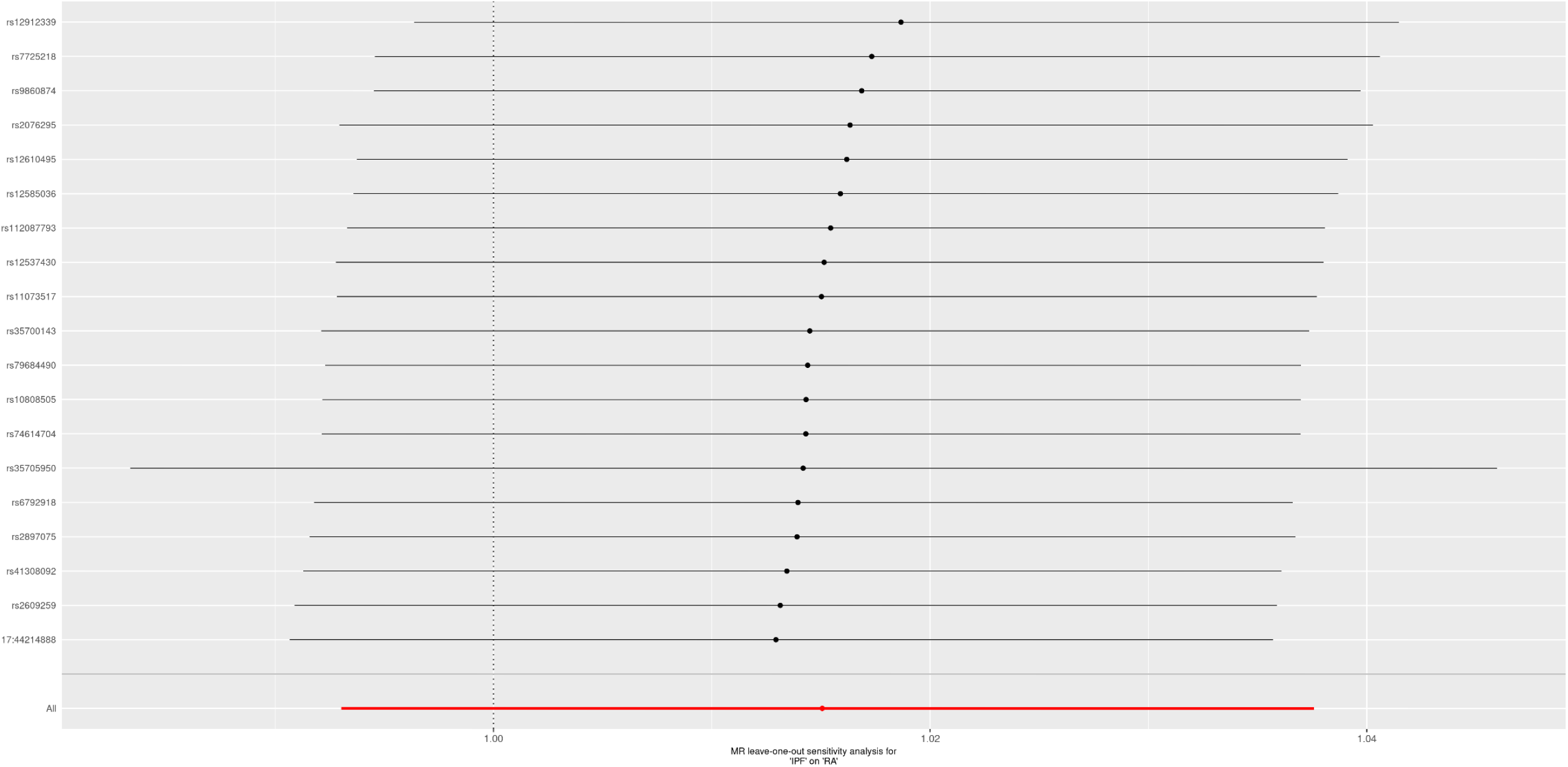
Leave one out analysis for estimation of causal effect of IPF on seronegative RA

## References

1. Olson AL, Swigris JJ, Sprunger DB, Fischer A, Fernandez-Perez ER, Solomon J, et al. Rheumatoid arthritis–interstitial lung disease–associated mortality. American journal of respiratory and critical care medicine. 2011;183(3):372–8.

2. Bongartz T, Nannini C, Medina-Velasquez YF, Achenbach SJ, Crowson CS, Ryu JH, et al. Incidence and mortality of interstitial lung disease in rheumatoid arthritis: a population-based study. Arthritis & Rheumatism. 2010;62(6):1583–91.

3. Koduri G, Norton S, Young A, Cox N, Davies P, Devlin J, et al. Interstitial lung disease has a poor prognosis in rheumatoid arthritis: results from an inception cohort. Rheumatology. 2010;49(8):1483–9.

4. Gabbay E, Tarala R, Will R, Carroll G, Adler B, Cameron D, et al. Interstitial lung disease in recent onset rheumatoid arthritis. American journal of respiratory and critical care medicine. 1997;156(2):528–35.

5. Hyldgaard C, Hilberg O, Pedersen AB, Ulrichsen SP, Løkke A, Bendstrup E, et al. A population-based cohort study of rheumatoid arthritis-associated interstitial lung disease: comorbidity and mortality. Ann Rheum Dis. 2017;76(10):1700–6.

6. Juge P, Lee JS, Lau J, Kawano-Dourado L, Serrano JR, Sebastiani M, et al. Methotrexate and rheumatoid arthritis associated interstitial lung disease. European Respiratory Journal. 2021;57(2).

7. Tanaka N, Kim JS, Newell JD, Brown KK, Cool CD, Meehan R, et al. Rheumatoid arthritis–related lung diseases: CT findings. Radiology. 2004;232(1):81–91.

8. Kelly CA, Saravanan V, Nisar M, Arthanari S, Woodhead FA, Price-Forbes AN, et al. Rheumatoid arthritis-related interstitial lung disease: associations, prognostic factors and physiological and radiological characteristics—a large multicentre UK study. Rheumatology. 2014;53(9):1676–82.

9. Assayag D, Lubin M, Lee JS, King TE, Collard HR, Ryerson CJ. Predictors of mortality in rheumatoid arthritis-related interstitial lung disease. Respirology. 2014;19(4):493–500.

10. Kim EJ, Elicker BM, Maldonado F, Webb WR, Ryu JH, Van Uden JH, et al. Usual interstitial pneumonia in rheumatoid arthritis-associated interstitial lung disease. European Respiratory Journal. 2010;35(6):1322–8.

11. Solomon JJ, Chung JH, Cosgrove GP, Demoruelle MK, Fernandez-Perez ER, Fischer A, et al. Predictors of mortality in rheumatoid arthritis-associated interstitial lung disease. European Respiratory Journal. 2016;47(2):588–96.

12. Kim EJ, Collard HR, King Jr TE. Rheumatoid arthritis-associated interstitial lung disease: the relevance of histopathologic and radiographic pattern. Chest. 2009;136(5):1397–405.

13. Matson S, Lee J, Eickelberg O. Two sides of the same coin? A review of the similarities and differences between idiopathic pulmonary fibrosis and rheumatoid arthritis-associated interstitial lung disease. European Respiratory Journal. 2021;57(5).

14. Solomon JJ, Danoff SK, Woodhead FA, Hurwitz S, Maurer R, Glaspole I, et al. Safety, tolerability, and efficacy of pirfenidone in patients with rheumatoid arthritis-associated interstitial lung disease: a randomised, double-blind, placebo-controlled, phase 2 study. The Lancet Respiratory Medicine. 2022.

15. Richeldi L, Du Bois RM, Raghu G, Azuma A, Brown KK, Costabel U, et al. Efficacy and safety of nintedanib in idiopathic pulmonary fibrosis. N Engl J Med. 2014;370(22):2071–82.

16. Juge P, Lee JS, Ebstein E, Furukawa H, Dobrinskikh E, Gazal S, et al. MUC5B promoter variant and rheumatoid arthritis with interstitial lung disease. N Engl J Med. 2018;379(23):2209–19.

17. Saag KG, Kolluri S, Koehnke RK, Georgou TA, Rachow JW, Hunninghake GW, et al. Rheumatoid arthritis lung disease. Determinants of radiographic and physiologic abnormalities. Arthritis & Rheumatism: Official Journal of the American College of Rheumatology. 1996;39(10):1711–9.

18. Baumgartner KB, Samet JM, Stidley CA, Colby TV, Waldron JA. Cigarette smoking: a risk factor for idiopathic pulmonary fibrosis. American journal of respiratory and critical care medicine. 1997;155(1):242–8.

19. Seibold MA, Wise AL, Speer MC, Steele MP, Brown KK, Loyd JE, et al. A common MUC5B promoter polymorphism and pulmonary fibrosis. N Engl J Med. 2011;364(16):1503–12.

20. Palomäki A, Palotie A, Koskela J, Eklund KK, Pirinen M, Ripatti S, et al. Lifetime risk of rheumatoid arthritis-associated interstitial lung disease in MUC5B mutation carriers. Ann Rheum Dis. 2021;80(12):1530–6.

21. Saevarsdottir S, Stefansdottir L, Sulem P, Thorleifsson G, Ferkingstad E, Rutsdottir G, et al. Multiomics analysis of rheumatoid arthritis yields sequence variants that have large effects on risk of the seropositive subset. Ann Rheum Dis. 2022.

22. Padyukov L. Genetics of rheumatoid arthritis. Seminars in Immunopathology; Springer; 2022.

23. Allen RJ, Stockwell A, Oldham JM, Guillen-Guio B, Schwartz DA, Maher TM, et al. Genome-wide association study across five cohorts identifies five novel loci associated with idiopathic pulmonary fibrosis. Thorax. 2022.

24. Myers TA, Chanock SJ, Machiela MJ. LDlinkR: an R package for rapidly calculating linkage disequilibrium statistics in diverse populations. Frontiers in genetics. 2020;11:157.

25. Staiger DO, Stock JH. Instrumental variables regression with weak instruments. Econometrica. 1994(65):557–586.

26. Lawlor DA, Harbord RM, Sterne JA, Timpson N, Davey Smith G. Mendelian randomization: using genes as instruments for making causal inferences in epidemiology. Stat Med. 2008;27(8):1133–63.

27. Burgess S, Butterworth A, Thompson SG. Mendelian randomization analysis with multiple genetic variants using summarized data. Genet Epidemiol. 2013;37(7):658–65.

28. Bowden J, Del Greco M F, Minelli C, Zhao Q, Lawlor DA, Sheehan NA, et al. Improving the accuracy of two-sample summary-data Mendelian randomization: moving beyond the NOME assumption. Int J Epidemiol. 2019;48(3):728–42.

29. Verbanck M, Chen C, Neale B, Do R. Detection of widespread horizontal pleiotropy in causal relationships inferred from Mendelian randomization between complex traits and diseases. Nat Genet. 2018;50(5):693–8.

30. Bowden J, Davey Smith G, Haycock PC, Burgess S. Consistent estimation in Mendelian randomization with some invalid instruments using a weighted median estimator. Genet Epidemiol. 2016;40(4):304–14.

31. Hartwig FP, Davey Smith G, Bowden J. Robust inference in summary data Mendelian randomization via the zero modal pleiotropy assumption. Int J Epidemiol. 2017;46(6):1985–98.

32. Bowden J, Davey Smith G, Burgess S. Mendelian randomization with invalid instruments: effect estimation and bias detection through Egger regression. Int J Epidemiol. 2015;44(2):512–25.

33. Bowden J, Del Greco M F, Minelli C, Davey Smith G, Sheehan NA, Thompson JR. Assessing the suitability of summary data for two-sample Mendelian randomization analyses using MR-Egger regression: the role of the I2 statistic. Int J Epidemiol. 2016;45(6):1961–74.

34. Kono M, Nakamura Y, Enomoto N, Hashimoto D, Fujisawa T, Inui N, et al. Usual interstitial pneumonia preceding collagen vascular disease: a retrospective case control study of patients initially diagnosed with idiopathic pulmonary fibrosis. PloS one. 2014;9(4):e94775.

35. Bernstein EJ, Barr RG, Austin JH, Kawut SM, Raghu G, Sell JL, et al. Rheumatoid arthritis-associated autoantibodies and subclinical interstitial lung disease: the Multi-Ethnic Study of Atherosclerosis. Thorax. 2016;71(12):1082–90.

36. Katsumata M, Hozumi H, Yasui H, Suzuki Y, Kono M, Karayama M, et al. Frequency and clinical relevance of anti-cyclic citrullinated peptide antibody in idiopathic interstitial pneumonias. Respir Med. 2019;154:102–8.

37. Mohning MP, Amigues I, Demoruelle MK, Pérez ERF, Huie TJ, Keith RK, et al. Duration of rheumatoid arthritis and the risk of developing interstitial lung disease. ERJ Open Research. 2021;7(1).

38. Samhouri BF, Vassallo R, Achenbach SJ, Kronzer VL, Davis III JM, Myasoedova E, et al. The Incidence, Risk Factors, and Mortality of Clinical and Subclinical Rheumatoid Arthritis-associated Interstitial Lung Disease: A Population-based Cohort. Arthritis Care & Research. 2022.

39. Solomon JJ, Matson S, Kelmenson LB, Chung JH, Hobbs SB, Rosas IO, et al. IgA antibodies directed against citrullinated protein antigens are elevated in patients with idiopathic pulmonary fibrosis. Chest. 2020;157(6):1513–21.

40. Holers VM, Demoruelle MK, Kuhn KA, Buckner JH, Robinson WH, Okamoto Y, et al. Rheumatoid arthritis and the mucosal origins hypothesis: protection turns to destruction. Nature Reviews Rheumatology. 2018;14(9):542–57.

41. Nielen MM, van Schaardenburg D, Reesink HW, Van de Stadt, Rob J, van der Horst-Bruinsma, Irene E, de Koning MH, et al. Specific autoantibodies precede the symptoms of rheumatoid arthritis: a study of serial measurements in blood donors. Arthritis & Rheumatism: Official Journal of the American College of Rheumatology. 2004;50(2):380–6.

42. Verpoort KN, Jol-Van Der Zijde, C M, Papendrecht-Van Der Voort E, Ioan-Facsinay A, Drijfhout JW, Van Tol M, et al. Isotype distribution of anti–cyclic citrullinated peptide antibodies in undifferentiated arthritis and rheumatoid arthritis reflects an ongoing immune response. Arthritis & Rheumatism: Official Journal of the American College of Rheumatology. 2006;54(12):3799–808.

43. Klareskog L, Stolt P, Lundberg K, Källberg H, Bengtsson C, Grunewald J, et al. A new model for an etiology of rheumatoid arthritis: smoking may trigger HLA–DR (shared epitope)–restricted immune reactions to autoantigens modified by citrullination. Arthritis & Rheumatism: Official Journal of the American College of Rheumatology. 2006;54(1):38–46.

44. Makrygiannakis D, Hermansson M, Ulfgren A, Nicholas AP, Zendman AJ, Eklund A, et al. Smoking increases peptidylarginine deiminase 2 enzyme expression in human lungs and increases citrullination in BAL cells. Ann Rheum Dis. 2008;67(10):1488–92.

45. Rangel-Moreno J, Hartson L, Navarro C, Gaxiola M, Selman M, Randall TD. Inducible bronchus-associated lymphoid tissue (iBALT) in patients with pulmonary complications of rheumatoid arthritis. J Clin Invest. 2006;116(12):3183–94.

46. Ytterberg AJ, Joshua V, Reynisdottir G, Tarasova NK, Rutishauser D, Ossipova E, et al. Shared immunological targets in the lungs and joints of patients with rheumatoid arthritis: identification and validation. Ann Rheum Dis. 2015;74(9):1772–7.

47. Giles JT, Danoff SK, Sokolove J, Wagner CA, Winchester R, Pappas DA, et al. Association of fine specificity and repertoire expansion of anticitrullinated peptide antibodies with rheumatoid arthritis associated interstitial lung disease. Ann Rheum Dis. 2014;73(8):1487–94.

48. Sokolove J, Bromberg R, Deane KD, Lahey LJ, Derber LA, Chandra PE, et al. Autoantibody epitope spreading in the pre-clinical phase predicts progression to rheumatoid arthritis. PloS one. 2012;7(5):e35296.

49. Konig MF, Abusleme L, Reinholdt J, Palmer RJ, Teles RP, Sampson K, et al. Aggregatibacter actinomycetemcomitans–induced hypercitrullination links periodontal infection to autoimmunity in rheumatoid arthritis. Science translational medicine. 2016;8(369):369ra176.

50. Engström M, Eriksson K, Lee L, Hermansson M, Johansson A, Nicholas AP, et al. Increased citrullination and expression of peptidylarginine deiminases independently of P. gingivalis and A. actinomycetemcomitans in gingival tissue of patients with periodontitis. Journal of translational medicine. 2018;16(1):1–14.

51. Invernizzi R, Wu BG, Barnett J, Ghai P, Kingston S, Hewitt RJ, et al. The respiratory microbiome in chronic hypersensitivity pneumonitis is distinct from that of idiopathic pulmonary fibrosis. American Journal of Respiratory and Critical Care Medicine. 2021;203(3):339–47.

52. Takahashi Y, Saito A, Chiba H, Kuronuma K, Ikeda K, Kobayashi T, et al. Impaired diversity of the lung microbiome predicts progression of idiopathic pulmonary fibrosis. Respiratory research. 2018;19(1):1–10.

53. Tong Y, Zheng L, Qing P, Zhao H, Li Y, Su L, et al. Oral microbiota perturbations are linked to high risk for rheumatoid arthritis. Frontiers in cellular and infection microbiology. 2020;9:475.

54. Cheng M, Zhao Y, Cui Y, Zhong C, Zha Y, Li S, et al. Stage-specific roles of microbial dysbiosis and metabolic disorders in rheumatoid arthritis. Ann Rheum Dis. 2022.

55. Burgess S, Smith GD, Davies NM, Dudbridge F, Gill D, Glymour MM, et al. Guidelines for performing Mendelian randomization investigations. Wellcome Open Research. 2019;4.

56. Kelly C, Emery P, Dieudé P. Current issues in rheumatoid arthritis-associated interstitial lung disease. The Lancet Rheumatology. 2021;3(11):e798–807.

57. Juge P, Granger B, Debray M, Ebstein E, Louis-Sidney F, Kedra J, et al. A Risk Score to Detect Subclinical Rheumatoid Arthritis-Associated Interstitial Lung Disease. Arthritis & Rheumatolog. 2022.

